# Development and Validation of a Simple Model to Predict Patient Height

**DOI:** 10.1101/2025.03.12.25323846

**Authors:** Emily E. Moin, Nicholas J. Seewald, Scott D. Halpern

**Affiliations:** Division of Pulmonary, Allergy, and Critical Care, University of Pennsylvania, Philadelphia; Palliative and Advanced Illness Research (PAIR) Center, University of Pennsylvania, Philadelphia; Department of Biostatistics, Epidemiology, and Informatics, University of Pennsylvania, Philadelphia

## Abstract

**Background:** Height recorded in electronic health records (EHRs) is used extensively in diagnosis and treatment, either in isolation or as a component of body-mass index (BMI), but is often falsely high because many adults overestimate their height. Statistical models to predict height could therefore improve population health, but to date models have required extensive input and have not been externally validated.

**Methods:** We used the National Health and Nutrition Examination Survey (NHANES) to develop sex-stratified predictive models for examiner-measured height based on self-reported height and age in a random 90% sample of data. We internally validated the model in a held-out 10% sample and externally validated the model in two cohorts: The National Adolescent to Adult Longitudinal Health Study (Add Health) and the University of Michigan Health and Retirement Study (HRS). We assessed discrimination with C-index, calibration by visual inspection of calibration plots, and accuracy using root mean square error (RMSE).

**Results:** Models were trained using 62,032 NHANES subjects (51.9% women, 21.7% Black, 23.9% Hispanic or Latino, with median age 48 [IQR 31 – 64]), and evaluated in the NHANES held-out test set (n=6,846), Add Health (n=5,749), and HRS (n=5,655). Models demonstrated excellent discrimination in all validation cohorts (C-index range 0.88 – 0.89). Models were well-calibrated in all validation cohorts. Model-predicted height demonstrated lower root mean square error (RMSE) compared to self-reported height in all validation cohorts and when stratified by race and ethnicity, with greatest improvements in participants aged 45 and over.

**Conclusions and Relevance:** A model requiring minimal input data improves estimation of height over self-reported height at least as much as more complex models across stratifications of sex, age, race and ethnicity in internal validation, and is the first model to improve height estimation that has demonstrated external validity.

---

Most adults self-report heights that are overestimates of their true height.^1–6^ Height is often obtained via self-report in clinical encounters, rather than formal stadiometry, leading to inaccurate information that is propagated throughout the electronic health record (EHR). Height alone is used for a broad spectrum of important diagnostic and therapeutic considerations, including diagnosis of obstructive and restrictive pulmonary disease with spirometry and specification of lung-protective ventilation in acute respiratory failure.^7,8^ Body mass index (BMI), a function of height and weight, is even more widely used, with applications in arenas as diverse as allocation of transplant organs,^9^ indications for medications like glucagon-like peptide-1 receptor agonists,^10,11^ and screening for risk of cardiovascular disease.^12^ Therefore, inaccurate height has wide-ranging consequences for individual medical care and population health.

Given the broad range of possible use cases for a more accurate estimate of adult height, and insufficient resources to measure height in all patient encounters, there is considerable opportunity to improve patient care with a broadly usable model to predict actual height. Prior attempts to develop techniques to adjust self-reported height and weight have relied on numerous input variables and either failed to undergo external validation^13,14^ or did not demonstrate external validity when evaluated.^15^ Thus, we sought to develop and externally validate a minimal model – i.e., one that relies on a small number of highly reliable input variables – that would be generalizable to the U.S. population and readily portable across health systems.

## Methods

### Model Development

We used data collected in the National Health and Nutrition Examination Survey (NHANES) to develop sex-stratified predictive linear regression models for examiner-measured height in adults (ages 18 and over).^16^ The University of Pennsylvania Institutional Review Board deemed this protocol exempt (#858042).

To maximize portability and generalizability, the sex-specific models’ inputs were limited to age and self-reported height. Because older ages are top-coded in NHANES for disclosure concerns, the top-coded ages were replaced with the average age in that group for each cohort. For example, in the NHANES 2021 – 2023 cohort, the average age of participants labeled with age 80 was 85, thus 85 was substituted in our dataset.

Sex-specific means and standard deviations were calculated for measured and self-reported height, and participants with values more than four standard deviations (SD) from their sex-specific mean in either measure were excluded. Four standard deviations was selected to facilitate model development while enabling predictions for an expected 99.9% of the population. Subjects with missing data for the main variables – age, sex, self-reported height, and examiner-measured height – were excluded from model development. Subjects with missing data for variables used for additional analyses (i.e., self-reported and examiner-measured weight, race and ethnicity) were included in model development and primary validation analyses but excluded from analyses requiring missing fields. No data were imputed.

We hypothesized a markedly non-linear relationship between self-reported and measured height based on inspection of histograms of self-reported height and knowledge that patients commonly use heuristics in estimating their height, portending over-reporting of certain heights (e.g., 72 inches or exactly 6 feet). Thus, we modeled self-reported height as a categorical variable with one level for each inch increment in height. Suspecting a non-linear relationship between age and measured height as well, we included age in our model as age strata spanning five years, except for the highest age stratum, which included all patients 85 and over. These were again included as separate categorical variables for flexibility.

Given the large size of the NHANES cohort, we used a fixed 90/10 training/validation split, with 90% of the cohort used to fit sex-specific linear regression models and 10% of the cohort held out for internal validation. To ensure identifiability of all height coefficients, the training/validation randomization was performed with stratification at the levels of height and sex.

Models were subsequently used to predict measured height for the held-out internal validation set and two external validation cohorts. The external validation cohorts are The National Longitudinal Adolescent to Adult Health Study (Add Health) Waves III, IV, and V^17^ and the University of Michigan Health and Retirement Study (HRS).^18^ Both Add Health and HRS collect self-reported and measured height, as well as age and sex, allowing for validation of our predictive model. For each cohort, we excluded age-sex strata with fewer than 10 participants. Because we included multiple waves of Add Health, we included only one observation per participant selected at random in the external validation dataset. Unlike NHANES, HRS records self-reported heights to the nearest 1/4”, thus self-reported heights were rounded to the nearest inch for model input. Records in both cohorts were excluded if they fell outside the 4 SD range as defined by NHANES, as the model is undefined outside this range.

### Statistical Analysis

Descriptive statistics were calculated for all included subjects. Predicted heights were calculated using the appropriate sex-specific model for each subject in the NHANES internal validation set and the two external validation sets. We used the standard formula^19^ to calculate five different versions of BMI for each participant: 1) measured BMI, based on examiner-measured height and examiner-measured weight; 2) self-reported BMI, based on self-reported height and self-reported weight; 3) clinical status quo (CSQ) BMI, based on self-reported height and examiner-measured weight; 4) model-predicted height with self-reported weight (hereafter referred to as model-self BMI), and 5) model-predicted height with examiner-measured weight (hereafter referred to as model-measured BMI).

We calculated the C-index for each cohort to assess discrimination. We assessed calibration visually using scatterplots of examiner-measured and model-predicted heights fit with locally estimated scatterplot smoothing (LOESS) curves. Mean absolute percent error (MAPE), root mean square error (RMSE), mean difference, and Bland-Altman limits of agreement (LOA) were calculated for each cohort and across relevant demographic stratifications to assess accuracy. To assess the added value of the model to the clinical status quo – i.e., using self-reported height as a proxy for measured height – we calculated these same metrics for self-reported heights in the validation sets and report the difference between our minimal model and the clinical status quo. Bootstrap standard errors were used to assess performance improvements for statistical significance and to calculate 95% confidence intervals (CI). Because many real world datasets do not specify whether provided heights are self-reported or examiner-measured, we compared performance of model-predicted heights when the input data are a random mixture of self-reported and examiner-measured heights, varying the proportions of these in the input data, in order to determine how many observations in a mixed-type dataset must be examiner-measured before model-adjustment fails to outperform naïve model inputs.

We used self-reported and examiner-measured weight in addition to height to assess the performance of our minimal model in two relevant binary classification tasks: 1) determination of normal weight (BMI < 25) vs. overweight or obese (BMI >= 25), and 2) determination of non-obese (BMI < 30) vs obese (BMI >= 30). Participants with missing data for self-reported or measured weight were excluded from these analyses. For binary classification tasks, we assessed performance using area under the receiver operating characteristic curve (AUROC) and Brier scores. To assess for marginal improvement of our minimal model over real-world scenarios, we calculate the scaled Brier score using the Brier score of BMI calculated with self-reported height as a reference, defined as 1 – model-predicted Brier score / reference Brier score, where 1 is best possible performance (i.e., 100% improvement over the reference approach) and the worst possible performance is an unbounded negative value. Analyses were performed using R version 4.4.2.

## Results

### Participants and Model Derivation

A total of 68,878 NHANES participants with complete data were included, with 62,032 (90.1%) assigned to the training set and 6,846 (9.9%) to internal validation. Models were fit using the training set (**Supplemental Table 1**). In the training set, 51.9% were women, 21.7% were Black, and 23.9% were Hispanic or Latino, with median age 48 (IQR 31 – 64). Based on examiner-measured height and weight, 36.5% were classified as obese, 32.6% as overweight, 29.0% as normal weight, and 1.9% as underweight. Similar characteristics were observed in the internal validation set. 5,749 Add Health participants (52.8% women, 22.2% Black, 10.7% Hispanic or Latino, median age 28 [IQR 23 – 35, range 18 – 42]) and 5,655 HRS participants (59.1% women, 38.3% Black, 28.1% Hispanic or Latino, median age 66 [IQR 59 – 74, range 40 – 101]) provided the two external validation cohorts (**Table 1**).

**Table 1.**
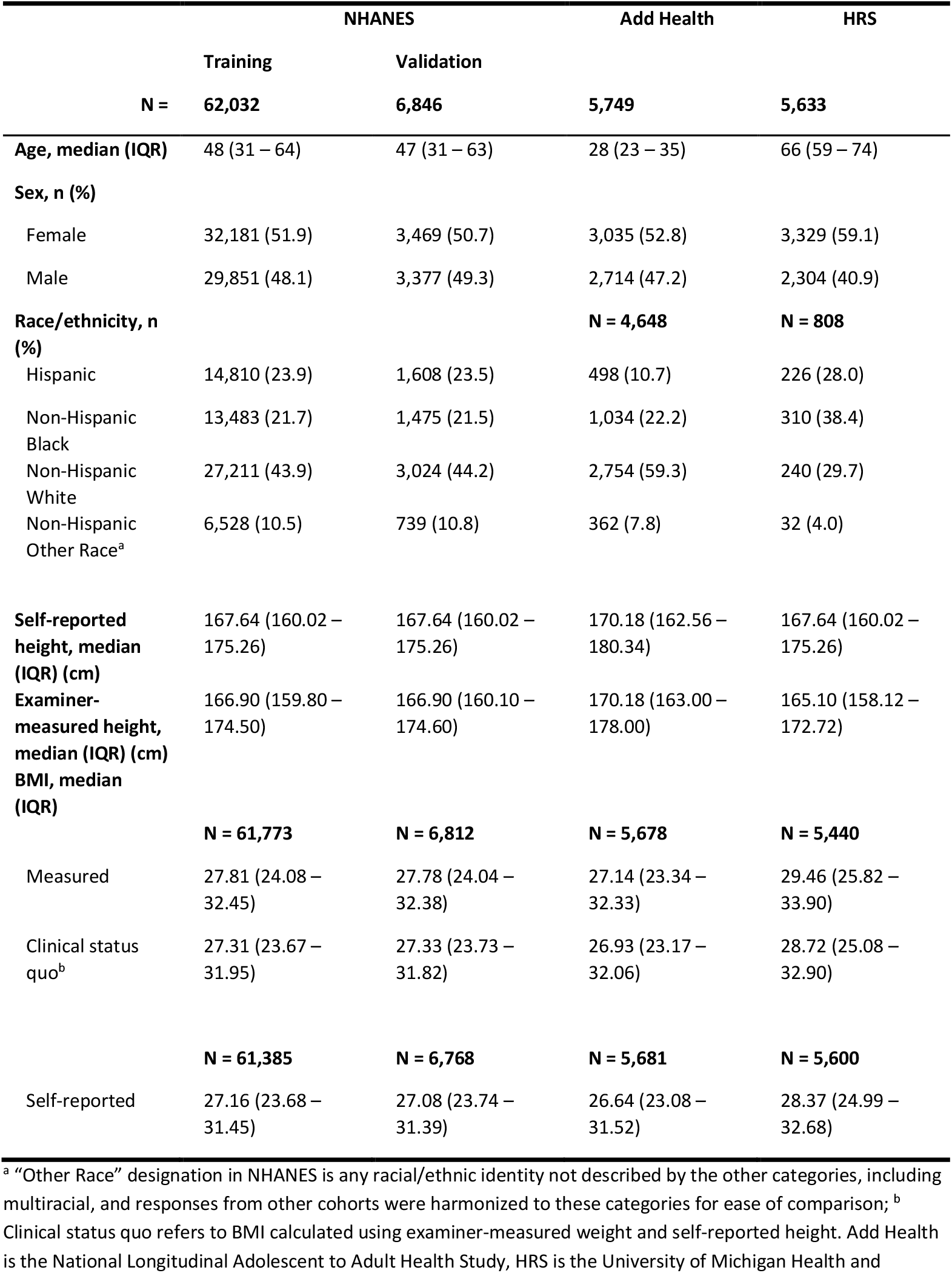
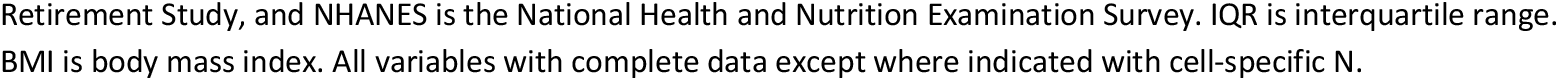
Participant characteristics by cohort.

|  | NHANES |  | Add Health | HRS |
| --- | --- | --- | --- | --- |
|  | Training | Validation |  |  |
| N = | 62,032 | 6,846 | 5,749 | 5,633 |
| **Age, median (IQR)** | 48 (31 – 64) | 47 (31 – 63) | 28 (23 – 35) | 66 (59 – 74) |
| **Sex, n (%)** |  |  |  |  |
| Female | 32,181 (51.9) | 3,469 (50.7) | 3,035 (52.8) | 3,329 (59.1) |
| Male | 29,851 (48.1) | 3,377 (49.3) | 2,714 (47.2) | 2,304 (40.9) |
| **Race/ethnicity, n (%)** |  |  | **N = 4,648** | **N = 808** |
| Hispanic | 14,810 (23.9) | 1,608 (23.5) | 498 (10.7) | 226 (28.0) |
| Non-Hispanic Black | 13,483 (21.7) | 1,475 (21.5) | 1,034 (22.2) | 310 (38.4) |
| Non-Hispanic White | 27,211 (43.9) | 3,024 (44.2) | 2,754 (59.3) | 240 (29.7) |
| Non-Hispanic Other Race<sup>a</sup> | 6,528 (10.5) | 739 (10.8) | 362 (7.8) | 32 (4.0) |
| **Self-reported height, median (IQR) (cm)** | 167.64 (160.02 – 175.26) | 167.64 (160.02 – 175.26) | 170.18 (162.56 – 180.34) | 167.64 (160.02 – 175.26) |
| **Examiner-measured height, median (IQR) (cm)** | 166.90 (159.80 – 174.50) | 166.90 (160.10 – 174.60) | 170.18 (163.00 – 178.00) | 165.10 (158.12 – 172.72) |
| **BMI, median (IQR)** | **N = 61,773** | **N = 6,812** | **N = 5,678** | **N = 5,440** |
| Measured | 27.81 (24.08 – 32.45) | 27.78 (24.04 – 32.38) | 27.14 (23.34 – 32.33) | 29.46 (25.82 – 33.90) |
| Clinical status quo<sup>b</sup> | 27.31 (23.67 – 31.95) | 27.33 (23.73 – 31.82) | 26.93 (23.17 – 32.06) | 28.72 (25.08 – 32.90) |
|  | **N = 61,385** | **N = 6,768** | **N = 5,681** | **N = 5,600** |
| Self-reported | 27.16 (23.68 – 31.45) | 27.08 (23.74 – 31.39) | 26.64 (23.08 – 31.52) | 28.37 (24.99 – 32.68) |

<sup>a</sup> “Other Race” designation in NHANES is any racial/ethnic identity not described by the other categories, including multiracial, and responses from other cohorts were harmonized to these categories for ease of comparison; <sup>b</sup> Clinical status quo refers to BMI calculated using examiner-measured weight and self-reported height. Add Health is the National Longitudinal Adolescent to Adult Health Study, HRS is the University of Michigan Health and
Retirement Study, and NHANES is the National Health and Nutrition Examination Survey. IQR is interquartile range. BMI is body mass index. All variables with complete data except where indicated with cell-specific N.

### Model Validation for Continuous Task

Discrimination was consistently good across all cohort-sex stratifications, with C-index values ranging from 0.880 – 0.893 for male participants and 0.876 – 0.884 for female participants. All models were well calibrated, as demonstrated in **Figure 1**.

**Figure 1.**
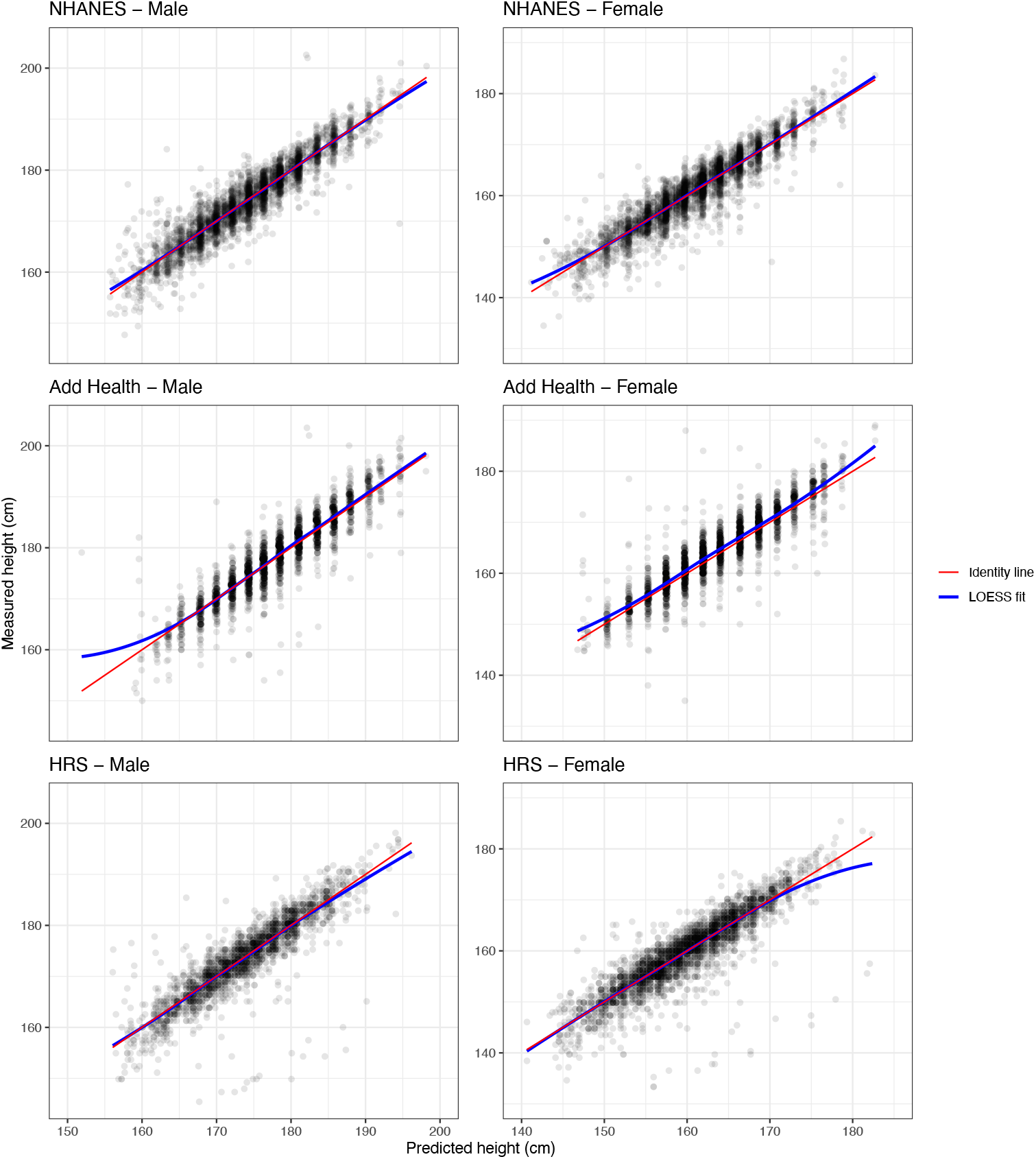
Calibration plots of predicted heights vs. measured heights stratified by cohort and sex. Each panel shows scatterplot of model-predicted height on x-axis and examiner-measured height on y-axis, with each translucent grey circle representing one observation in that cohort. Red line is the identity line (predicted height exactly equal to measured height) and blue line is a LOESS (locally estimated scatterplot smoothing) fit, such that perfect overlap of the identity line and LOESS fit line would indicate perfect model calibration.

RMSE improvement, defined as the difference between RMSE of model-predicted heights and RMSE of self-reported heights, demonstrated statistically significant superior model performance in all cohort-sex stratifications. The greatest improvement in RMSE was observed in male HRS participants (1.03 cm difference, 95% CI 0.95 – 1.12), the smallest in female Add Health participants (0.11 cm difference, 95% CI 0.03 – 0.20) (**Table 2**).

**Table 2.** Root mean square error (RMSE) stratified by cohort, sex, and race/ethnicity.

| Race/ethnicity | Sex | Cohort | Self-reported | Root mean square error (cm) |  |  |
| --- | --- | --- | --- | --- | --- | --- |
|  |  |  |  | Model-predicted | Improvement | 95% CI |
| All race/ethnicities | Male | NHANES | 3.56 | 2.80 | 0.76 | 0.67 – 0.85 |
|  |  | Add | 3.41 | 3.14 | 0.27 | 0.17 – 0.35 |
|  |  | HRS | 4.72 | 3.69 | 1.03 | 0.95 – 1.12 |
|  | Female | NHANES | 3.24 | 2.71 | 0.54 | 0.46 – 0.62 |
|  |  | Add | 3.15 | 3.04 | 0.11 | 0.03 – 0.20 |
|  |  | HRS | 4.14 | 3.33 | 0.82 | 0.74 – 0.92 |
| Hispanic | Male | NHANES | 4.04 | 3.35 | 0.69 | 0.54 – 0.87 |
|  |  | Add | 3.18 | 3.00 | 0.17 | 0 – 0.34 |
|  |  | HRS | 6.50 | 5.29 | 1.21 | 0.80 – 1.61 |
|  | Female | NHANES | 4.22 | 3.47 | 0.75 | 0.60 – 0.93 |
|  |  | Add | 3.90 | 3.50 | 0.40 | -0.03 – 1.06 |
|  |  | HRS | 6.41 | 5.43 | 0.99 | 0.58 – 1.40 |
| Non-Hispanic White | Male | NHANES | 3.52 | 2.57 | 0.95 | 0.78 – 1.07 |
|  |  | Add | 3.16 | 2.96 | 0.20 | 0.03 – 0.32 |
|  |  | HRS | 4.07 | 3.16 | 0.91 | 0.61 – 1.24 |
|  | Female | NHANES | 2.68 | 2.29 | 0.39 | 0.29 – 0.50 |
|  |  | Add | 2.78 | 2.74 | 0.04 | -0.04 – 0.11 |
|  |  | HRS | 2.61 | 2.06 | 0.55 | 0.21 – 0.88 |
| Non-Hispanic Black | Male | NHANES | 3.32 | 2.80 | 0.52 | 0.25 – 0.74 |
|  |  | Add | 3.35 | 3.29 | 0.06 | -0.32 – 0.35 |
|  |  | HRS | 5.81 | 4.81 | 1.00 | 0.60 – 1.35 |
|  | Female | NHANES | 3.25 | 2.72 | 0.54 | 0.40 – 0.66 |
|  |  | Add | 3.56 | 3.49 | 0.07 | -0.05 – 0.19 |
|  |  | HRS | 4.16 | 3.35 | 0.80 | 0.61 – 1.10 |
| Other Race | Male | NHANES | 3.06 | 2.39 | 0.67 | 0.51 – 0.81 |
|  |  | Add | 4.28 | 3.89 | 0.39 | -0.01 – 0.73 |
|  |  | HRS | 3.41 | 3.26 | 0.15 | -1.51 – 1.35 |
|  | Female | NHANES | 2.77 | 2.29 | 0.48 | 0.28 – 0.75 |
|  |  | Add | 3.02 | 2.95 | 0.07 | -0.12 – 0.29 |
|  |  | HRS | 3.54 | 2.69 | 0.85 | 0 – 1.60 |

Add Health is the National Longitudinal Adolescent to Adult Health Study, HRS is the University of Michigan Health and Retirement Study, and NHANES is the National Health and Nutrition Examination Survey. 95% confidence intervals (CI) calculated using bootstrapping.

When stratified by race and ethnicity in addition to sex, model-predicted RMSE point estimates were improved compared to self-reported height RMSE for all stratifications in all cohorts, but with reduced power. The greatest improvement in RMSE over self-reported height was observed for Hispanic male participants in HRS (1.21 cm difference, 95% CI 0.80 – 1.61), and the least for Non-Hispanic White female participants in Add Health (0.04 cm difference, 95% CI −0.04 – 0.11) (**Table 2**). MAPE, mean difference, and LOA demonstrated similar trends across sex, cohort, and race/ethnicity stratifications (**Supplemental Tables 2 and 3**).

When stratified by age and sex, model-predicted RMSE demonstrated consistently good performance, with RMSE ranging from 2.2 cm in female participants aged 40 – 44 in HRS and male participants aged 30 – 34 in NHANES to 5.8 cm in male participants aged 85 and over in HRS. Compared to self-reported RMSE, model predictions outperformed self-report for all age, sex, and cohort stratifications except Add Health males aged 20 – 24, Add Health females aged 35 – 44, and HRS females aged 40 – 44, though none of these differences were statistically significant. RMSE improvements were generally greater with increasing age, with progressively greater improvements over self-report observed in participants aged 45 and over (**Figure 2**).

**Figure 2.**
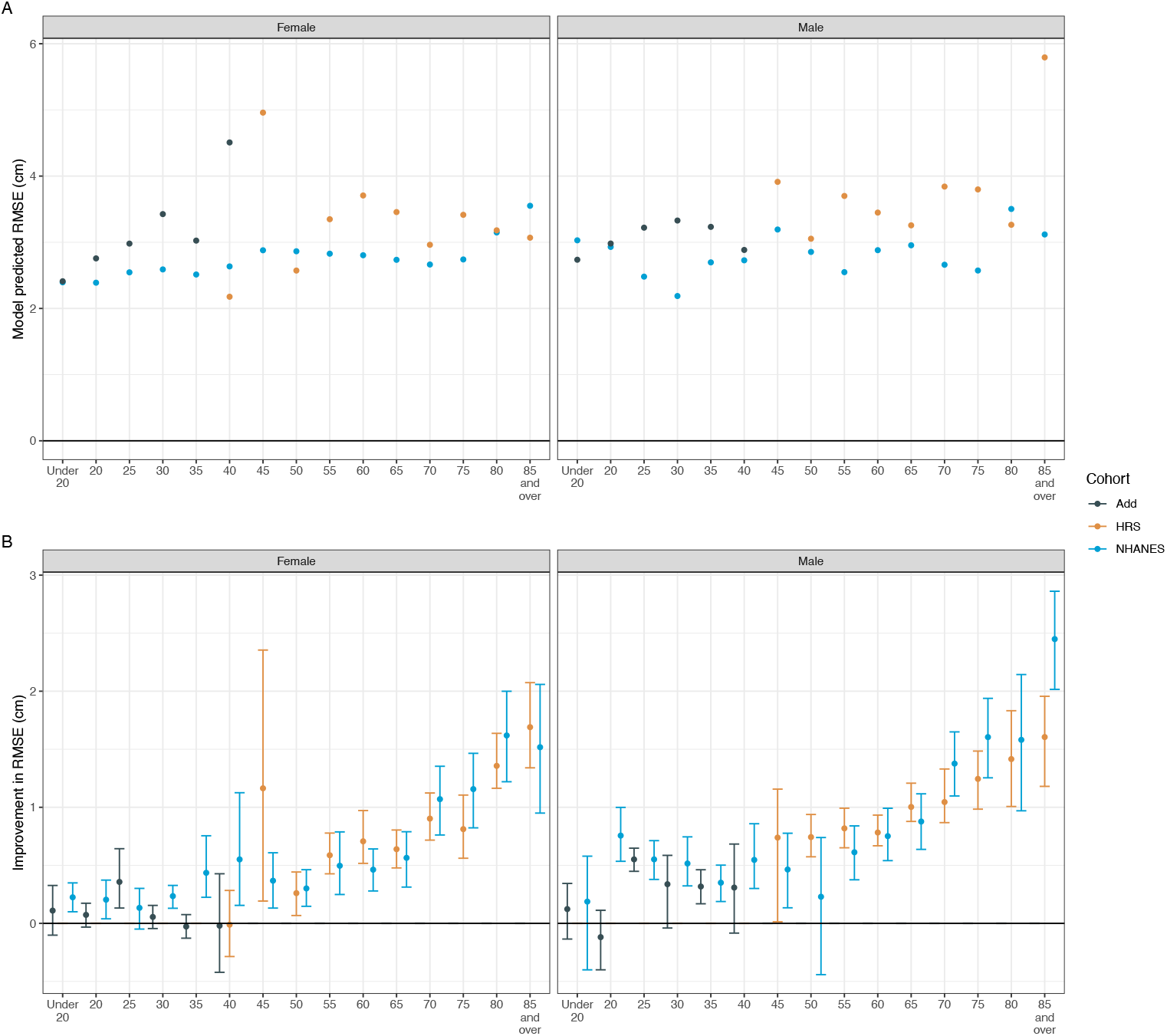
A. Model predicted RMSE stratified by age, sex, and cohort. B. Improvement in RMSE of model-predicted over self-reported RMSE by age, sex, and cohort. X-axes of all panels display age in 5-year bins. RMSE is root mean square error. Add is the National Longitudinal Adolescent to Adult Health Study, HRS is the University of Michigan Health and Retirement Study, and NHANES is the National Health and Nutrition Examination Survey. Model predicted RMSE (panel A) represents a comparison between examiner-measured heights (gold standard) and a predicted height based on age, sex, and self-reported height. In panel B, improvement in RMSE is the difference between model predicted RMSE and the RMSE calculated by comparing self-reported height to examiner-measured heights. Error bars represent 95% CI calculated with bootstrapping. All RMSE are reported in centimeters.

When model input data were randomized to mixtures of self-reported and examiner-measured heights, model adjustment yielded improved RMSE over model inputs despite large amounts of admixed examiner-measured heights across both sexes in all cohorts. Across stratifications of cohort and sex, performance among female participants in NHANES was least sensitive to mixed input data, with model adjustment leading to improved performance when input data contained less than 64.3% examiner-measured data. Performance among female participants in Add Health was the most sensitive to mixed input data, such that performance gains were realized by the model only if 32.7% or fewer estimates represented examiner-measured data (**Figure 3**).

**Figure 3.**
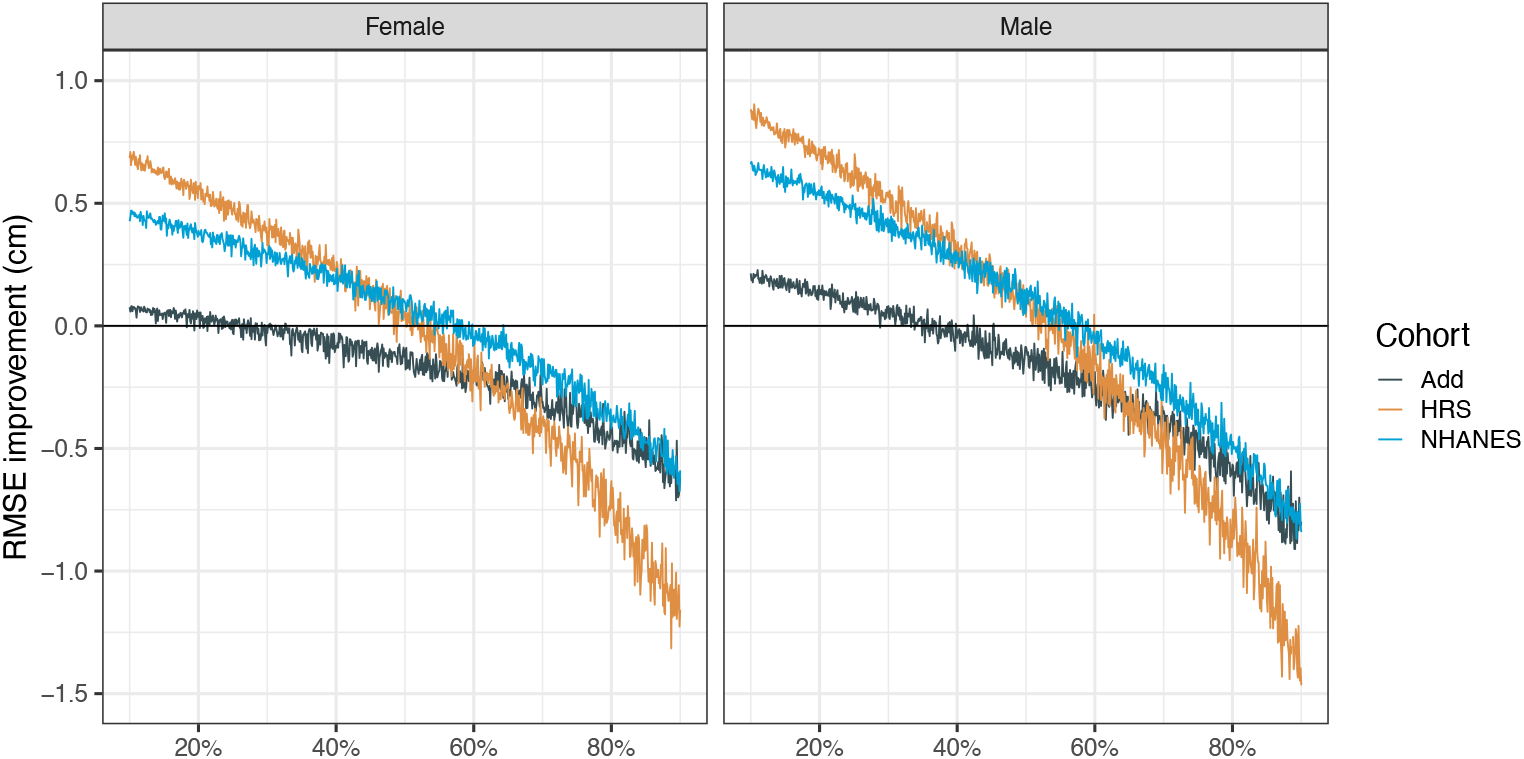
Improvement in RMSE after model-adjustment as a function of percentage of height observations that are examiner-measured instead of self-reported, stratified by sex and cohort. X-axis represents percentage of input data that are examiner-measured heights (as opposed to self-reported heights). Y-axis is improvement of model-predicted height over input data represented as the difference in RMSE. Horizontal solid line at zero represents equal performance between model-predicted heights and no model prediction. RMSE is root mean square error. Add is the National Longitudinal Adolescent to Adult Health Study, HRS is the University of Michigan Health and Retirement Study, and NHANES is the National Health and Nutrition Examination Survey. Improvement in RMSE is the difference between model predicted RMSE (model-predicted height compared to gold-standard examiner-measured height) and the RMSE calculated by comparing self-reported height to examiner-measured heights. For each datapoint, a percentage of observations (given by “correctly labeled observations” on the x-axis) were randomized to have their input data be self-reported height and the remainder to be examiner-measured height. All observations with RMSE improvement greater than zero represent improved performance when using model-adjustment despite some amount of data coming from examiner-measurement in the input dataset. All RMSE are reported in centimeters.

### Model Validation for Classification Task

AUROC for classification of BMI >= 25 and BMI >= 30 ranged from 0.986 – 0.995 when model-predicted height was combined with examiner-measured weight (model-measured BMI) and 0.967 – 0.985 when combining model-predicted height with self-reported weight (model-self BMI) across sex and cohort stratifications (**Supplemental Table 4**). Using model-predicted height instead of self-reported height improved AUROC across all cutoffs, BMI types, sexes, and cohorts, except for Add Health male participants. Brier scores for BMI classification were less than 0.06 for all cutoffs, sexes, and cohorts for model-measured BMI, and less than 0.09 for model-self BMI.

Scaled Brier scores were calculated relative to a reference Brier score for BMI classification based on self-reported height (i.e., percent improvement in Brier score when model-predicted height was substituted for self-reported height). When comparing model-measured BMI to CSQ BMI, scaled Brier scores demonstrated improvement with model-predicted height across all cutoffs, BMI types, sexes, and cohorts, apart from Add Health, where performance decrement ranged from −8.3% to 0.0%. Performance improvements in other groups ranged from 0.8% to 31.9% (**Supplemental Table 5**).

When examining changes in BMI classification at the individual level, incorporating model-adjusted height into BMI calculations resulted in reclassification from an incorrect to a correct BMI category for 0.6 – 5.0% of participants across cohorts, sexes, and classification tasks. There was also reclassification from a correct to incorrect BMI category for 0.5 – 2.1% of participants. Correct reclassifications were greater than incorrect reclassifications in all cohorts except Add Health, where net gain of correctly classified participants ranged from −0.7% to 0.3% (**Supplemental Table 6**).

## Discussion

In this study, we report a novel model for the prediction of true (examiner-measured) height based on self-reported height, sex, and age. We show that this minimal model using linear regression achieves excellent performance in predicting examiner-measured height, demonstrates improved performance over commonly used self-reported height, and results in improved BMI classification over formulae utilizing self-reported height. Moreover, we show that the model still improves performance when input data include a mix of both self-reported and examiner-measured heights, as is common in clinical datasets.

Discrepancy between self-reported and measured height has previously been shown to be associated with increasing age,^1–6^ likely owing to the infrequent practice of formal height measurement in adults and a tendency for adults to lose height as they age. Consistent with this, although our model demonstrates good performance in height prediction across the range of adult ages, it provides the greatest performance gains over self-reported height in patients aged 45 and older. Correspondingly, the more limited improvements in performance among younger patients likely reflects a ceiling effect – because self-reported height is a better estimate of measured height in younger adults, it is more difficult to improve upon it.

Our model surpasses prior efforts to estimate true height in three important ways: First, several previous attempts^13,14,20^ have developed models with extensive input variables, such as education level, geographic region, or race. By including only age and sex as additional covariates, our model is more widely applicable and avoids the problematic biologization of race that has been heavily scrutinized in diagnostic and predictive algorithms.^21–24^ Furthermore, there is increasing recognition of significant measurement error even in self-reported race and ethnicity,^25^ further increasing the desirability of a model whose performance does not rely upon these variables. Indeed, despite not including race or ethnicity as input variables, our model nevertheless performs well across all race and ethnicity stratifications in our validation cohorts.

Second, previous models have either not been validated externally^13,14^ or, upon external validation, were found to lack predictive validity.^15^ Our model demonstrated improved performance over self-report in a held-out internal validation set as well as two external cohorts. Moreover, the internal validation metrics of our model match or exceed those of previous models that were only internally validated. For example, An and Ji developed a variety of machine learning models for predicting height and weight using age, sex, race and ethnicity as input variables, and report RMSE by performing 10-fold cross-validation of their training data.^14^ Out of all architectures tested, they achieved a best RMSE of 2.8 cm using the XGBoost model architecture. Our linear regression model’s RMSE in our held-out internal validation set was 2.8 cm and 2.7 cm in males and females, respectively, and this performance was maintained in external cohorts without the use of race or ethnicity input data. Scholes et al. developed models for height and weight prediction exclusively using linear regression with stepwise selection of variables for model parsimony.^13^ Among their tested models, they reported a best mean difference of −0.02 cm in males and −0.10 cm in females when tested against a held-out internal validation set. By comparison, our model achieved equivalent performance in males (0.02 cm) and superior performance in females (−0.06 cm) when evaluated in our held-out internal validation set.

Finally, our simple model using linear regression requires minimal computing resources while still achieving similar or better performance to models using more sophisticated machine learning methods.^14^

## Limitations

A notable limitation of our model is that while it demonstrated consistently good performance and improvement over self-report across cohorts and relevant stratifications, its use less consistently improved classification of BMI among individuals. Specifically, although using the model produced a net gain in correct classifications over what would be achieved through self-report alone, its use shifted some participants from a correct to incorrect classification. Thus, prior to deploying the model in a specific clinical context (e.g., determining treatment eligibility based on a BMI cutoff), a randomized trial or other experimental study should examine overall outcomes among patients managed with vs. without the model.

## Conclusion

The use of self-reported height as a surrogate for true, examiner-measured height is systematically biased. Instead, a simple model to estimate examiner-measured height based on three easily accessible inputs consistently improves model calibration and discrimination compared with self-report across subgroups defined by age, sex, race and ethnicity.

## Data Availability

All data produced in the present study are available upon reasonable request to the authors.

https://www.cdc.gov/nchs/nhanes/index.html

https://hrs.isr.umich.edu/about

https://addhealth.cpc.unc.edu/

## Acknowledgments

This research uses data from Add Health, funded by grant P01 HD31921 (Harris) from the *Eunice Kennedy Shriver* National Institute of Child Health and Human Development (NICHD), with cooperative funding from 23 other federal agencies and foundations. Add Health is currently directed by Robert A. Hummer and funded by the National Institute on Aging cooperative agreements U01 AG071448 (Hummer) and U01AG071450 (Aiello and Hummer) at the University of North Carolina at Chapel Hill. Add Health was designed by J. Richard Udry, Peter S. Bearman, and Kathleen Mullan Harris at the University of North Carolina at Chapel Hill. The HRS (Health and Retirement Study) is sponsored by the National Institute on Aging (grant number NIA U01AG009740) and is conducted by the University of Michigan. This analysis uses Early Release data from the Health and Retirement Study, 2022 Core Early Release (Version 2.0)), sponsored by the National Institute on Aging (grant number NIA U01AG009740) and conducted by the University of Michigan. These data have not been cleaned and may contain errors that will be corrected in the Final Public Release version of the dataset.

## References

1. Hodge JM, Shah R, McCullough ML, Gapstur SM, Patel AV. Validation of self-reported height and weight in a large, nationwide cohort of U.S. adults. PLOS ONE. 2020;15(4):e0231229. doi:10.1371/journal.pone.0231229

2. Connor Gorber S, Tremblay M, Moher D, Gorber B. A comparison of direct vs. self-report measures for assessing height, weight and body mass index: a systematic review. Obes Rev Off J Int Assoc Study Obes. 2007;8(4):307–326. doi:10.1111/j.1467-789X.2007.00347.x

3. Maukonen M, Männistö S, Tolonen H. A comparison of measured versus self-reported anthropometrics for assessing obesity in adults: a literature review. Scand J Public Health. 2018;46(5):565–579. doi:10.1177/1403494818761971

4. Niedhammer I, Bugel I, Bonenfant S, Goldberg M, Leclerc A. Validity of self-reported weight and height in the French GAZEL cohort. Int J Obes Relat Metab Disord J Int Assoc Study Obes. 2000;24(9):1111–1118. doi:10.1038/sj.ijo.0801375

5. Spencer EA, Appleby PN, Davey GK, Key TJ. Validity of self-reported height and weight in 4808 EPIC-Oxford participants. Public Health Nutr. 2002;5(4):561–565. doi:10.1079/PHN2001322

6. Taylor AW, Dal Grande E, Gill TK, et al. How valid are self-reported height and weight? A comparison between CATI self-report and clinic measurements using a large cohort study. Aust N Z J Public Health. 2006;30(3):238–246. doi:10.1111/j.1467-842x.2006.tb00864.x

7. The Acute Respiratory Distress Syndrome Network. Ventilation with Lower Tidal Volumes as Compared with Traditional Tidal Volumes for Acute Lung Injury and the Acute Respiratory Distress Syndrome. N Engl J Med. 2000;342(18):1301–1308. doi:10.1056/NEJM200005043421801

8. Agustí A, Celli BR, Criner GJ, et al. Global Initiative for Chronic Obstructive Lung Disease 2023 Report: GOLD Executive Summary. Eur Respir J. 2023;61(4):2300239. doi:10.1183/13993003.00239-2023

9. Kidney transplant candidacy evaluation and waitlisting practices in the US and their association with access to transplantation - PMC. Accessed February 19, 2025. https://pmc.ncbi.nlm.nih.gov/articles/PMC9177783/

10. Wilding JPH, Batterham RL, Calanna S, et al. Once-Weekly Semaglutide in Adults with Overweight or Obesity. N Engl J Med. 2021;384(11):989–1002. doi:10.1056/NEJMoa2032183

11. Jastreboff AM, Roux CW le, Stefanski A, et al. Tirzepatide for Obesity Treatment and Diabetes Prevention. N Engl J Med. 0(0). doi:10.1056/NEJMoa2410819

12. 2013 AHA/ACC/TOS guideline for the management of overweight and obesity in adults: a report of the American College of Cardiology/American Heart Association Task Force on Practice Guidelines and The Obesity Society - PubMed. Accessed February 19, 2025. https://pubmed.ncbi.nlm.nih.gov/24222017/

13. Scholes S, Ng Fat L, Moody A, Mindell JS. Does the use of prediction equations to correct self-reported height and weight improve obesity prevalence estimates? A pooled cross-sectional analysis of Health Survey for England data. BMJ Open. 2023;13(1):e061809. doi:10.1136/bmjopen-2022-061809

14. An R, Ji M. Building Machine Learning Models to Correct Self-Reported Anthropometric Measures. J Public Health Manag Pract. 2023;29(5):671. doi:10.1097/PHH.0000000000001769

15. Flegal KM, Graubard BI, Ioannidis JPA. Evaluation of a suggested novel method to adjust BMI calculated from self-reported weight and height for measurement error. Obes Silver Spring Md. 2021;29(10):1700–1707. doi:10.1002/oby.23239

16. CDC. National Health and Nutrition Examination Survey. National Health and Nutrition Examination Survey. January 15, 2025. Accessed January 23, 2025. https://www.cdc.gov/nchs/nhanes/index.html

17. Harris KM, Halpern CT, Whitsel EA, et al. Cohort Profile: The National Longitudinal Study of Adolescent to Adult Health (Add Health). Int J Epidemiol. 2019;48(5):1415–1415k. doi:10.1093/ije/dyz115

18. Produced and distributed by the University of Michigan with funding from the National Institute on Aging (grant number NIA U01AG009740). Health and Retirement Study, (2022 Core Early Release (Version 2.0)) public use dataset. Published online September 2024.

19. Keys A, Fidanza F, Karvonen MJ, Kimura N, Taylor HL. Indices of relative weight and obesity. J Chronic Dis. 1972;25(6):329–343. doi:10.1016/0021-9681(72)90027-6

20. Ward ZJ, Bleich SN, Cradock AL, et al. Projected U.S. State-Level Prevalence of Adult Obesity and Severe Obesity. N Engl J Med. 2019;381(25):2440–2450. doi:10.1056/NEJMsa1909301

21. Vyas DA, Eisenstein LG, Jones DS. Hidden in Plain Sight — Reconsidering the Use of Race Correction in Clinical Algorithms. N Engl J Med. 2020;383(9):874–882. doi:10.1056/NEJMms2004740

22. Vyas DA, Jones DS, Meadows AR, Diouf K, Nour NM, Schantz-Dunn J. Challenging the Use of Race in the Vaginal Birth after Cesarean Section Calculator. Womens Health Issues. 2019;29(3):201–204. doi:10.1016/j.whi.2019.04.007

23. Vasan RS, Heuvel E van den. Differences in estimates for 10-year risk of cardiovascular disease in Black versus White individuals with identical risk factor profiles using pooled cohort equations: an in silico cohort study. Lancet Digit Health. 2022;4(1):e55–e63. doi:10.1016/S2589-7500(21)00236-3

24. Schluger NW, Dozor AJ, Jung YEG. Rethinking the Race Adjustment in Pulmonary Function Testing. Ann Am Thorac Soc. 2022;19(3):353–356. doi:10.1513/AnnalsATS.202107-890PS

25. Salhi RA, Macy ML, Samuels-Kalow ME, Hogikyan M, Kocher KE. Frequency of Discordant Documentation of Patient Race and Ethnicity. JAMA Netw Open. 2024;7(3):e240549. doi:10.1001/jamanetworkopen.2024.0549

